# COVID-19 Modelling: the Effects of Social Distancing

**DOI:** 10.1101/2020.03.29.20046870

**Authors:** Oliva Bendtsen Cano, Sabrina Cano Morales, Claus Bendtsen

## Abstract

The purpose of this article is to reach all those who find it difficult to become well informed about the repercussions of a lockdown strategy to tackle the COVID-19 pandemic and to spark discussion and thought. Here we use simple stochastic simulations to evaluate different approaches taken to tackle the crisis, along with the efficiency they will hold and the number of casualties they may incur. It is clear that the less strict the social distancing the more time it will take for life to return to normal, and the more lives will be at risk. This is shown through simulations formed by an open sourced code, which allows evaluation of the outcomes from different intervention scenarios or conditions.

## Introduction

As we know, different countries have taken different approaches to try and keep the pandemic at bay; with China going on complete lockdown, the UK only going on semi-lockdown with people still going to work, and Sweden not taking any particular approach to locking down, the most efficient method is unclear. It is hard for everyone to attain the necessary information, and the information is usually rather difficult for non-experts to understand.

So, this article attempts to give people the necessary information about the changing aspects of the COVID-19 pandemic from a scientific perspective but in a simple manner, using only elementary level mathematics. Here, we make the necessary tools available to undergo ‘experiments’ at home; open sourced code is provided in order for people to see the outcomes of certain actions by the formation of graphs or summary statistics. We hope that using this material will help readers put numbers shared on the news into context.

Another goal that this article aims to achieve is to make people understand the consequences of their own actions when it comes to this pandemic – everybody has to do their bit, so it is crucial to have everybody understand what it is that they exactly have to do.

### Epidemiological and clinical observations

Since the outbreak of the COVID-19 virus, a number of studies trying to explain the dynamics of the outbreak have been published. These studies draw mainly from publicly available data of cases reported by hospitals, WHO, China Centre for Control Disease and other health care organisations.

There seems to be consensus regarding the incubation period, which has been reported with a mean of 5.0 days [1], 6.4 days [2], 5.5 days [3]. It seems reasonable to assume this to be about 5.5 days perhaps with a range of 2 to 14 days, also consistent with the conclusions in [4]. However, as reported by Xu [5] incubation appears to be longer in tertiary patients, which will have an effect in our UK population.

From the time when symptoms appear to the patient’s hospitalisation (in those cases when needed) the median is of 7.0 days [2], or as reported by Li [1] between 3.3 and 6.5 days depending on the outcome.

Fatality rate of patients varies depending on whether we are looking at overall cases or only those of hospitalised patients. Mortality for all known cases vary from 2% and 3% [6] to 3.5% [2], while if looking at hospitalised patients, the number goes up to 7% [7], 4% to 11% [6], 8.2% [4] and even reaches 13.9% [8]. Demographic is a sizable variable in these rates, reaching up to 50%–75% for the elderly and those patients with comorbidities [6]. Health care systems, as well, are major players in the outcome of hospitalised patients.

Basic reproduction number or *R*_0_ has been reported between 2 and 3.5 [2], 2.2 and 3.6 [9], and 2 and 6.5 [6].

Finally, the number of asymptomatic cases is difficult to estimate. Three cases of group isolation have allowed for some models to appear and asymptomatic rate has been reported at 17.9% [10], 30% or less than a half [11] and between 50% and 75% as reported by Prof. Romagnani from the University of Florence [12].

## Methods

### A Markov Chain Model for COVID-19

We use a very simply Markov chain model to represent the dynamics of the epidemic. In Figure 1 it is illustrated how individuals can move between states. Starting from being healthy, they move to becoming infected, then to shedding the virus (i.e. being contagious) before becoming symptomatic. From symptomatic they can become sick (which we use to represent hospitalised) and may die. The recovery back to healthy (represented as immune, which seems plausible [13], although it is not yet well-understood if reinfections can occur) can happen from either the shedding, symptomatic or sick state.

**Figure 1:**
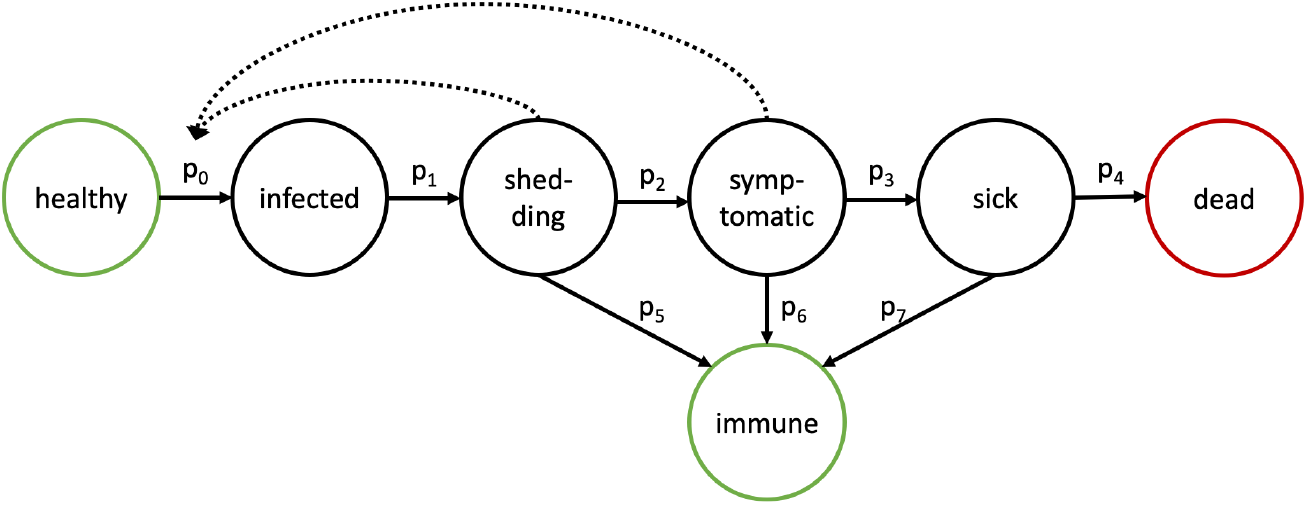
A Markov Chain model describing how individuals can transition between states after infection.

People in the shedding and symptomatic state infect healthy people a rate proportional to how many they are.

We model the transition probabilities, *p*_1_ to *p*_7_ following an Erlang dis-tribution, represented as *ℰ* but scaled when multiple outcomes from a state is possible. This allows us to mimic observed dynamics of when people move from one state to another.

The transition probability, *p*_0_ can be represented through a desired basic reproduction number, *R*_0_ as it can be easily shown that:

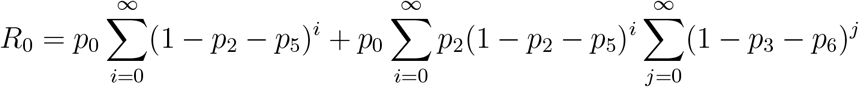

To mimic the epidemiological and clinial observations summarised in the introduction we use *R*_0_ = 2.75, *p*_1_ *∼ ℰ* (10, 4), *p*_2_ *∼* (1 *− w*_5_) *ℰ* (3, 1.5), *p*_3_ *∼* (1 *− w*_6_) *ℰ* (14, 2), and *p*_4_ *∼* (1 *− w*_7_) *ℰ* (4, 1), We empirically consider the recovery distributions and fix these as *p*_5_ *∼ w*_5_ *ℰ* (5, 0.5), *p*_6_ *∼ w*_6_ *ℰ* (10, 1), and *p*_7_ *w*_7_ (6, 1) with *w*_5_ = 0.85, *w*_6_ = 0.9, and *w*_7_ = 0.95.

R [14] was used to implement the model and a markdown [15] implementation of all results is available as supplementary information.

## Results

### Model Validation

To assess how well the model recapitulates what we currently know about COVID-19 we conduct a simulation of the basic model and investigate the distributions of time between events, as shown in Figure 2. We observe a time from infection to becoming symptomatic between approximately 2 and 14 days with a mode around day 6. Time from symptomatic to sick is observed to peak at 7 to 8 days and time from hospitalisation to death varies between 1 and 10 days approximately.

**Figure 2:**
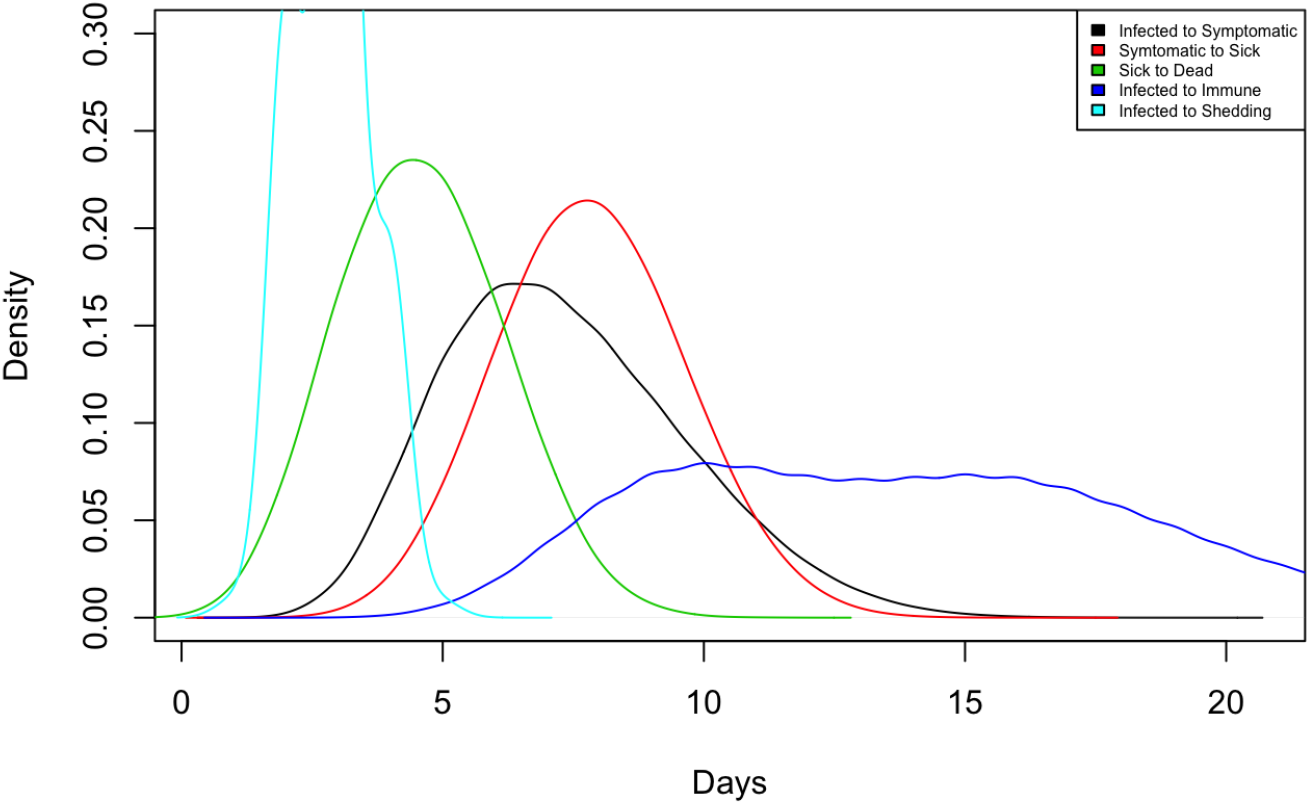
Observed densities of time between events in the model.

To compare the initial trajectories for mortalities and hospitalised with what has been observed in Italy and the United Kingdom, we overlay data for either country as shown in Figure 3.

**Figure 3:**
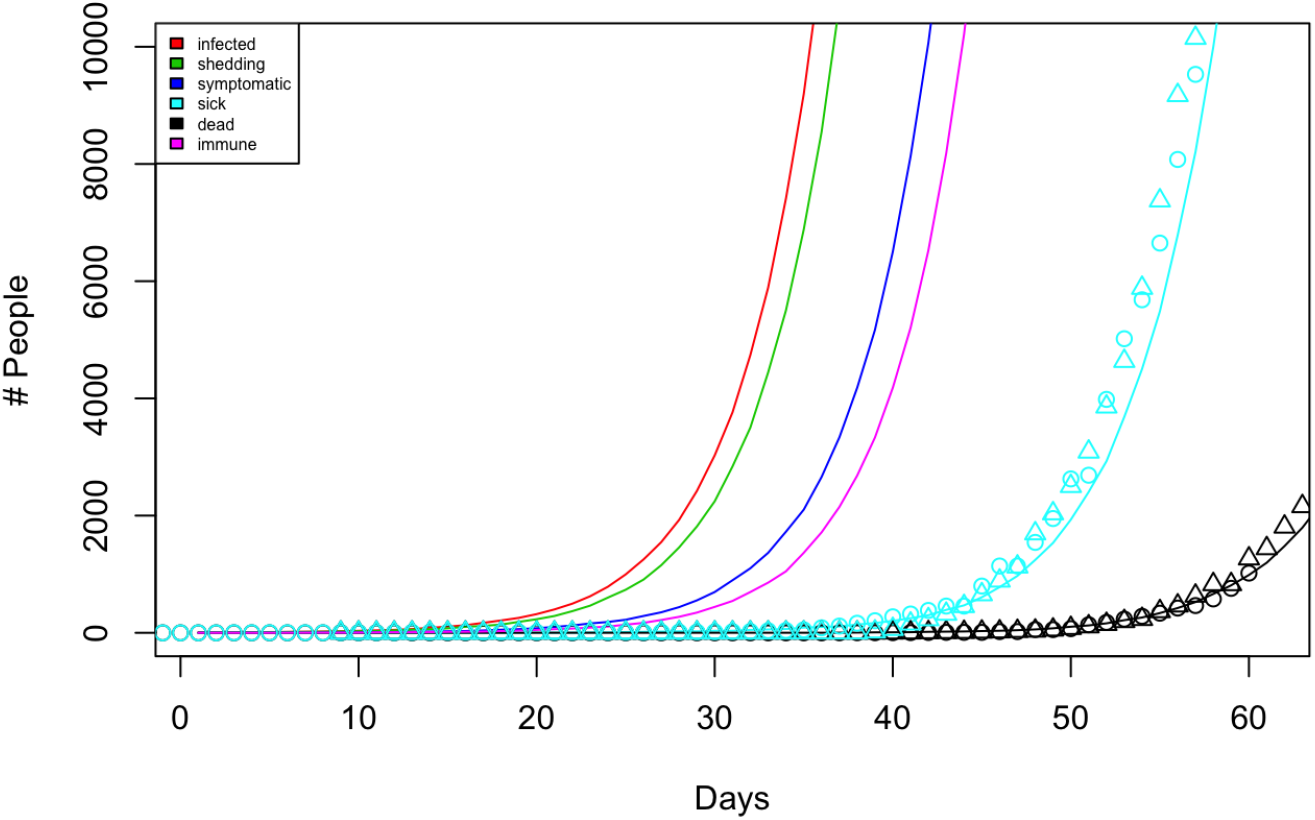
Model simulations overlaid with observational data of reported cases and mortalities from Italy (△) and the United Kingdom (*◦*).

### Simulating the UK

To understand the effect of the virus at population scale, we simulate the effect of a constant *R*_0_ = 2.75 for a population of 60 million which is seeded by 10 infected individuals on day 1. The results are shown on Figure 4 and we observe an overall duration of the epidemic of 149 days, with the number of people sick peaking on day 88. The overall mortality rate is 1.32%.

**Figure 4:**
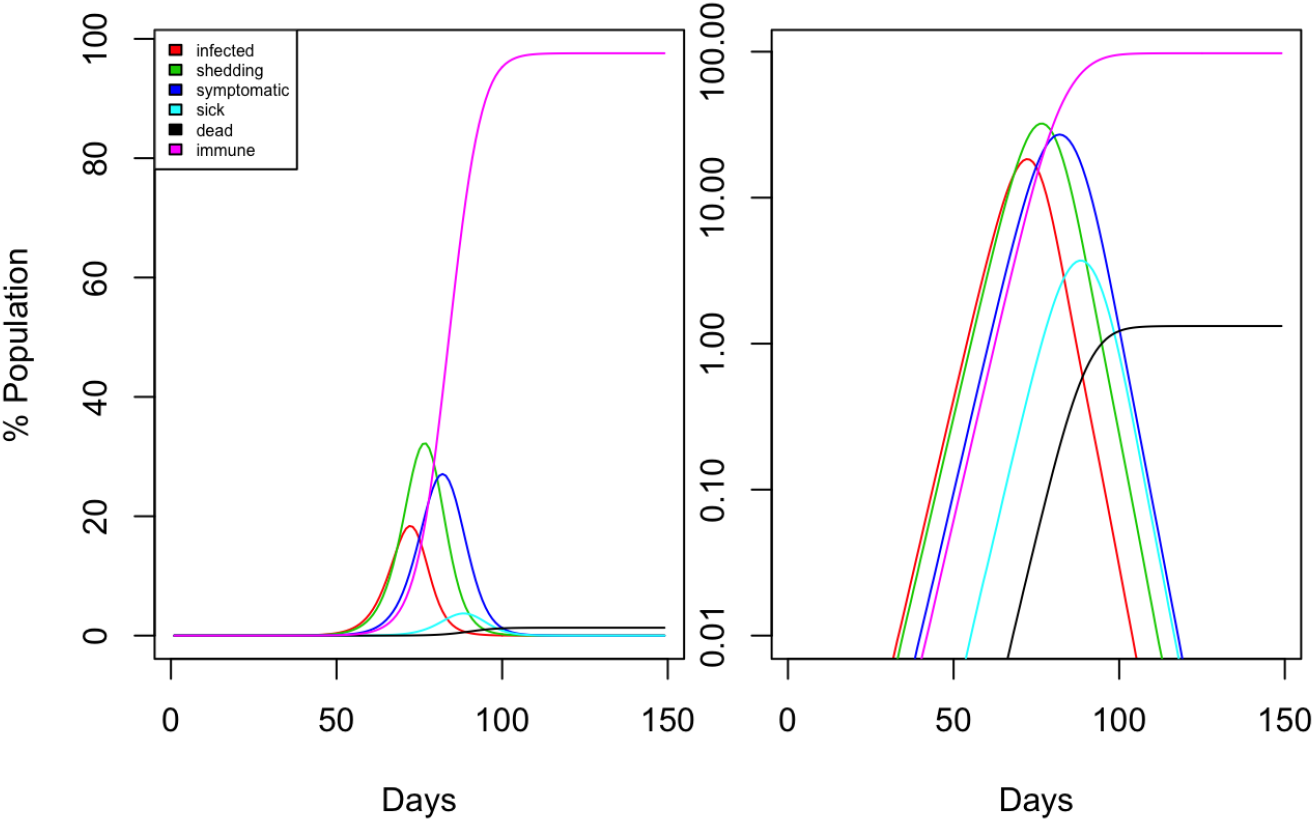
Model simulations with constant *R*_0_ = 2.75 and a population size of 60 million.

Next, we consider the effect of social distancing. This can readily be implemented in the model simply by redefining the basic reproduction number, *R*_0_ at the day the measures are implemented to reflect a sufficient decline in how many people each shedding or symptomatic person will infect. On one extreme we have perfect social distancing with *R*_0_ = 0 but we also investigate the effect of *R*_0_ = 0.5 and *R* = 0.75.

Day 55 is used for initiating social distancing since the simulations here showed 337 dead and 5486 sick consistent with reported numbers on 23 March 2020 when the United Kingdom initiated lockdown.

In the case of perfect social distancing the mortality rate is only 0.04% (21474 dead) and the epidemic is resolved by day 90 with the number of people sick peaking on day 70.

In the case of a more relaxed social distancing with *R*_0_ = 0.50 the mortality rate is 0.13% (79781 dead) without having the epidemic resolved by day 250 and with the number of people sick peaking on day 71. A somewhat stricter social distancing with *R*_0_ = 0.25 would resolve the epidemic by day 189 and lead to 32998 dead.

Finally, if the social distancing is relaxed to *R*_0_ = 0.75 we observe a much later peak in the number of people sick on day 112 and also a much larger mortality rate of 0.55% (330964 dead).

## Discussion

As can be seen from Figure 2, the simulations displayed above correlate well with data found in the biomedical literature. As seen in Figure 3, the number of deaths in Italy and the United Kingdom correlate well with the predicted deaths. The number of recorded cases also conform well with the simulation, however they may be slightly higher due to the larger number of people getting tested than actually being hospitalised. All in all, it is clearly seen that the simulation overall reflects the available data well.

Now, looking at the simulations for the UK, we can explore the different outcomes for the different approaches taken. If, like Sweden, the UK were to take the route of relying on herd immunity, as seen in Figure 4, almost all of the population becomes immune (with the exception of the 1.3% of the population that die). The length of time that this will take is just over three months as seen in the graphs. So, the result of this approach would be a relatively fast outcome with a large percentage of people immune, but with the downside of having an estimated 800,000 mortalities.

However, the UK has not taken this approach and has instead decided to adopt a social distancing approach and go on lockdown. If UK’s lockdown goes perfectly (as shown in Figure 5), the pandemic will be resolved in roughly months with only 21, 000 dead. However, seeing as UK is only going on a semi-lockdown (with e.g. many people still going to work and using public transportation), the likelihood of this seems very low. It seems much more likely that the result of this semi-lockdown looks more like Figure 6, with 4.5 months of doing this semi-lockdown and with approximately 80, 000 dead.

**Figure 5:**
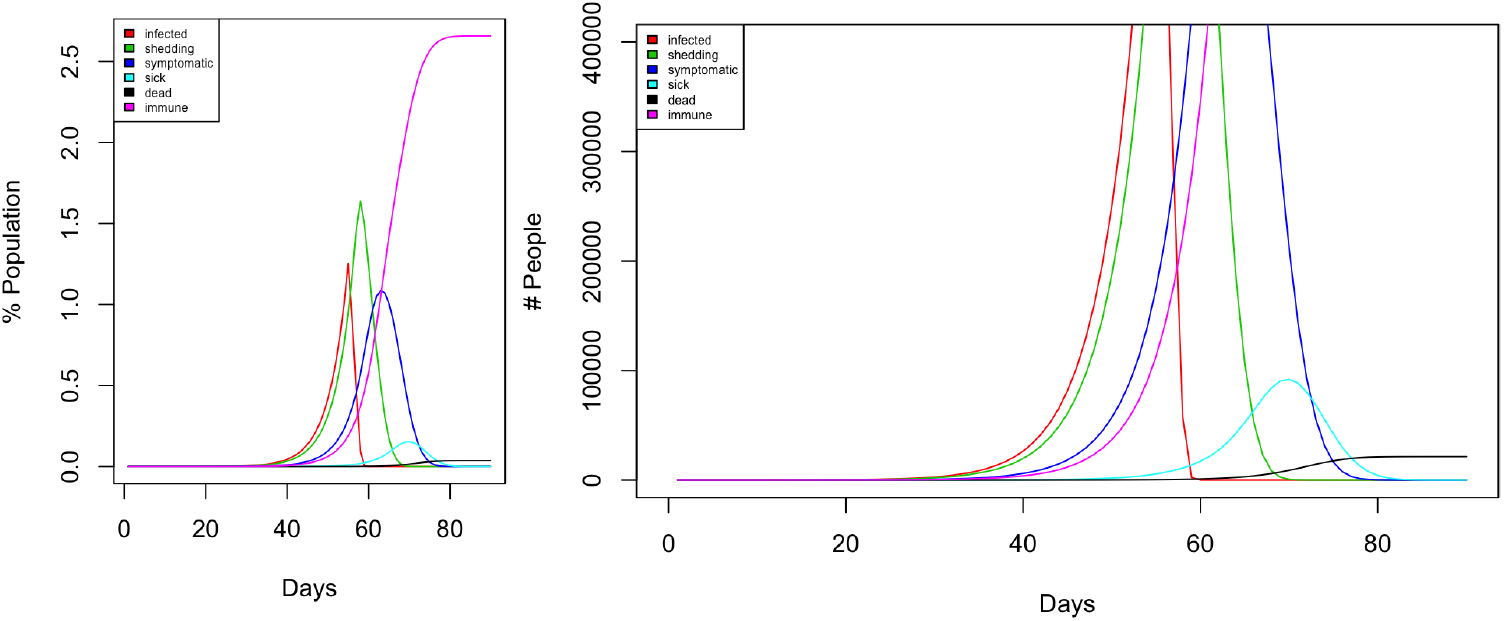
Model simulations implementing perfect social distancing with *R*_0_ = 0 from day 55 for a population size of 60 million.

**Figure 6:**
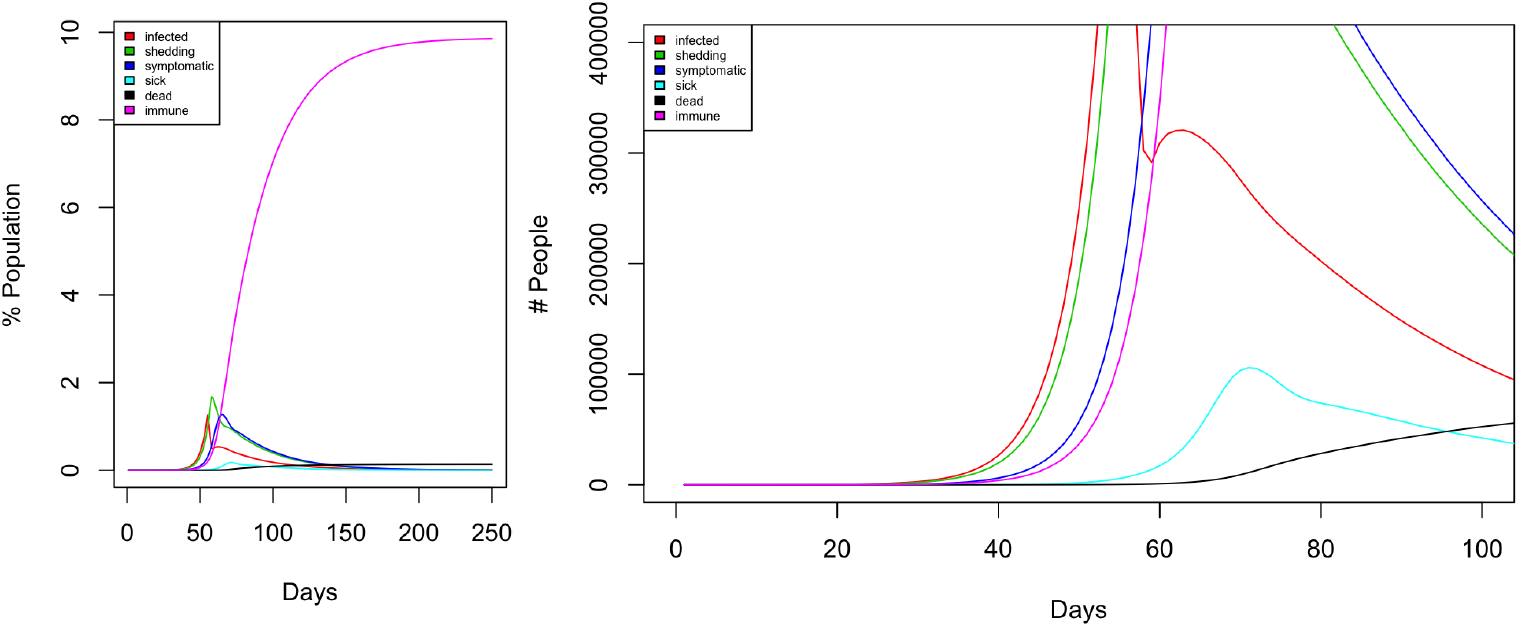
Model simulations implementing social distancing with *R*_0_ = 0.50 from day 55 for a population size of 60 million.

We can also see what happens if the lockdown is even more relaxed, with people going out at times that aren’t completely necessary because people start to grow bored. If people do start to take this lockdown less seriously — especially young people who believe that they will not get gravely ill due to their youth – it might result in things looking more like Figure 7, with the ‘lockdown’ taking over 6+ months and resulting in more than 300, 000 fatalities. This reflects just how important it is to stay at home; the more relaxed we become the longer this is going to take with more deaths occurring, which is what no one wants.

**Figure 7:**
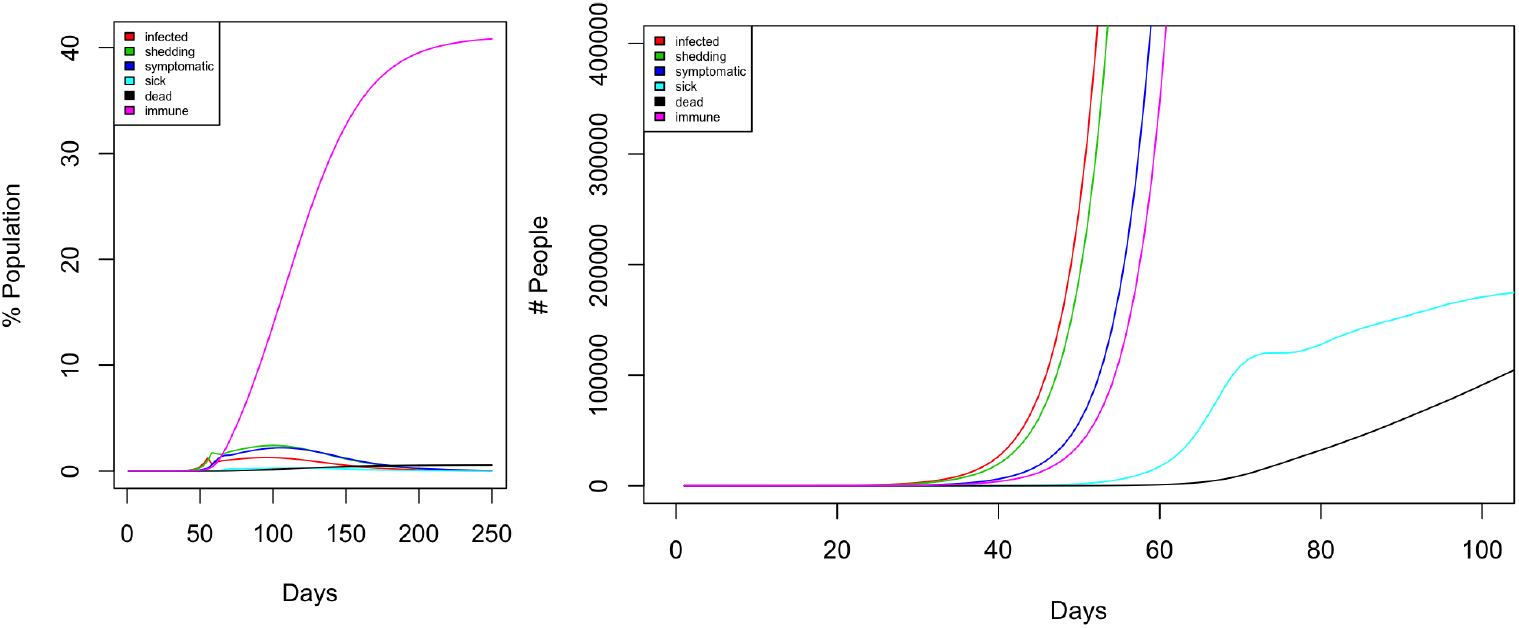
Model simulations implementing social distancing with *R*_0_ = 0.75 from day 55 for a population size of 60 million.

To conclude on the simulations, we can see that the less seriously lock-down is taken, or the less strict we are about this lockdown, the longer this will take. If this lockdown isn’t done to a high enough standard, it is evident that this will take much longer, with the results not looking too different to that of taking the herd immunity approach.

We have made the code available to everyone to use. We encourage people to use it and to look at the consequence of different national or international actions. If, for example, one would like to test and see what would happen if people start to relax after one month of strict lockdown, this can simply be done by increase the *R*_0_ number at that point and evaluate the results. This is one of the advantages from using a simply stochastic simulation approach as presented herein, as opposed to more classical approaches in epidemiological modelling based on differential equations [16].

Finally, one caveat. This is just a model (and all models are wrong [17]) – further validation and stress testing, including sensitivity analysis of the results to the choice of parameters should be done to understand the limitations of the model predictions.

## Data Availability

all data to reproduce the results in the manuscript are included as supplementary material

## References

[1] Natalie M Linton, Tetsuro Kobayashi, Yichi Yang, Katsuma Hayashi, Andrei R Akhmetzhanov, Sung-mok Jung, Baoyin Yuan, Ryo Kinoshita, and Hiroshi Nishiura. Incubation Period and Other Epidemiological Characteristics of 2019 Novel Coronavirus Infections with Right Truncation: A Statistical Analysis of Publicly Available Case Data. Journal of Clinical Medicine, 9(2):538–9, February 2020.

[2] Yixuan Wang, Yuyi Wang, Yan Chen, and Qingsong Qin. Unique epidemiological and clinical features of the emerging 2019 novel coronavirus pneumonia (COVID-19) implicate special control measures. Journal of Medical Virology, pages jmv.25748–28, March 2020.

[3] Stephen A Lauer, Kyra H Grantz, Qifang Bi, Forrest K Jones, Qulu Zheng, Hannah R Meredith, Andrew S Azman, Nicholas G Reich, and Justin Lessler. The Incubation Period of Coronavirus Disease 2019 (COVID-19) From Publicly Reported Confirmed Cases: Estimation and Application. Annals of internal medicine, pages 1–7, March 2020.

[4] Chih-Cheng Lai, Yen Hung Liu, Cheng-Yi Wang, Ya-Hui Wang, Shun-Chung Hsueh, Muh-Yen Yen, Wen-Chien Ko, and Po-Ren Hsueh. Asymptomatic carrier state, acute respiratory disease, and pneumonia due to severe acute respiratory syndrome coronavirus 2 (SARS-CoV-2): Facts and myths. Journal of Microbiology, Immunology and Infection, pages 1–9, March 2020.

[5] Tianmin Xu, Cong Chen, Zhen Zhu, Manman Cui, Chunhua Chen, Hong Dai, and Yuan Xue. Clinical features and dynamics of viral load in imported and non-imported patients with COVID-19. International Journal of Infectious Diseases, pages 1–18, March 2020.

[6] Tanu Singhal. A Review of Coronavirus Disease-2019 (COVID-19). Indian journal of pediatrics, 87(4):281–286, April 2020.

[7] Long quan Li, Tian Huang, Yong qing Wang, Zheng ping Wang, Yuan Liang, Tao bi Huang, Hui yun Zhang, Wei ming Sun, and Yu ping Wang. 2019 novel coronavirus patients’ clinical characteristics, discharge rate and fatality rate of meta-analysis. Journal of Medical Virology, pages jmv.25757–7, March 2020.

[8] Alfonso J Rodriguez-Morales, Jaime A Cardona-Ospina, Estefanea Gutierrez-Ocampo, Rhuvi Villamizar-Peña, Yeimer Holguin-Rivera, Juan Pablo Escalera-Antezana, Lucia Elena Alvarado-Arnez, D Katterine Bonilla-Aldana, Carlos Franco-Paredes, Andres F Henao-Martinez, Alberto Paniz-Mondolfi, Guillermo J Lagos-Grisales, Eduardo Ramerez-Vallejo, Jose A Suerez, Lysien I Zambrano, Wilmere Villamil-Gómez, Graciela J Balbin-Ramon, Ali A Rabaan, Harapan Harapan, Kuldeep Dhama, Hiroshi Nishiura, Hiromitsu Kataoka, Tauseef Ahmad, Ranjit Sah, and Latin American Network of Coronavirus Disease 2019-COVID-19 Research LANCOVID-19. Clinical, laboratory and imaging features of COVID-19: A systematic review and meta-analysis. Travel Medicine and Infectious Disease, page 101623, March 2020.

[9] Shi Zhao, Qianyin Lin, Jinjun Ran, Salihu S Musa, Guangpu Yang, Weiming Wang, Yijun Lou, Daozhou Gao, Lin Yang, Daihai He, and Maggie H Wang. Preliminary estimation of the basic reproduction number of novel coronavirus (2019-nCoV) in China, from 2019 to 2020: A data-driven analysis in the early phase of the outbreak. International Journal of Infectious Diseases, 92:214–217, March 2020.

[10] Kenji Mizumoto, Katsushi Kagaya, Alexander Zarebski, and Gerardo Chowell. Estimating the asymptomatic proportion of coronavirus disease 2019 (COVID-19) cases on board the Diamond Princess cruise ship, Yokohama, Japan, 2020. Euro surveillance : bulletin Europeen sur les maladies transmissibles = European communicable disease bulletin, 25(10):454, March 2020.

[11] Hiroshi Nishiura, Tetsuro Kobayashi, Ayako Suzuki, Sung-mok Jung, Katsuma Hayashi, Ryo Kinoshita, Yichi Yang, Baoyin Yuan, Andrei R Akhmetzhanov, Natalie M Linton, and Takeshi Miyama. Estimation of the asymptomatic ratio of novel coronavirus infections (COVID-19). International Journal of Infectious Diseases, pages 1–9, March 2020.

[12] Michael Day. Covid-19: identifying and isolating asymptomatic people helped eliminate virus in Italian village. BMJ, pages 1–1, March 2020.

[13] Irani Thevarajan, Thi H O Nguyen, Marios Koutsakos, Julian Druce, Leon Caly, Carolienae Sandt, Xiaoxiao Jia, Suellen Nicholson, Mike Catton, Benjamin Cowie, Steven Y C Tong, Sharon R Lewin, and Katherine Kedzierska. Breadth of concomitant immune responses prior to patient recovery: a case report of non-severe COVID-19. Nature Medicine, pages 1–10, March 2020.

[14] R Core Team. R: A Language and Environment for Statistical Computing. R Foundation for Statistical Computing, Vienna, Austria, 2019.

[15] Yihui Xie, J.J. Allaire, and Garrett Grolemund. R Markdown: The Definitive Guide. Chapman and Hall/CRC, Boca Raton, Florida, 2018. ISBN 9781138359338.

[16] Herbert W Hethcote. Three basic epidemiological models. In Applied mathematical ecology, pages 119–144. Springer, 1989.

[17] Gep Box. Science and Statistics. Journal of the American Statistical Association, 71, 1976.

